# Cycle threshold values in symptomatic COVID-19 cases in England

**DOI:** 10.1101/2022.06.13.22276321

**Authors:** Sebastian Funk, Sam Abbott

## Abstract

**Introduction:** Since the start of the pandemic SARS-CoV-2 infection has most commonly been confirmed using reverse transcriptase polymerase chain reaction (RT-PCR), with results translated into a binary positive/negative outcomes. Previous studies have found that there is additional useful information in the level of the Cycle threshold (Ct value) of positive cases. Here we characterise variation in Ct values as a proxy for viral loads in more than 3 million test-positive COVID-19 cases in England with the aim of better quantifying the utility of such data.

**Methods:** We used individual N gene Ct values from symptomatic PCR positive (with Ct value less than 30) Pillar 2 cases in England who self-reported the date of symptom onset, and for whom age, reinfection status, variant status, and the number of vaccines received was available. Those with a positive test result more than 6 days after their reported symptom onset were excluded to mitigate the potential impact of recall bias. We used a generalised additive model, to estimate Ct values empirical mean Ct values for each strata of interest independently as well as to predict Ct values using a model that adjusted for a range of demographic and epidemiological covariates jointly. We present empirical Ct values and compare them to predicted mean Ct values.

**Results:** We found that mean Ct values varied by vaccine status, and reinfection status with the number of vaccine doses having little apparent effect. Modelling Ct values as a smooth function of time since onset and other variables struggled to reproduce the individual variation in the data but did match the population-level variation over time relatively well with this being apparently dominated by large differences between variants. Other variation over time was also captured to some degree though their remained several periods where the model could not capture the empirical means with a potential explanation being epidemic phase bias.

**Conclusions:** Analysing a large dataset of routine Ct values from symptomatic COVID-19 cases in England we found variation based on time since symptom onset, vaccine status, age, and variant. Ct values were highest 1-3 days after symptom onset and differed most due to variant status. We found no clear correlation between previously estimated differences in intrinsic transmissibility and Ct values indicating that this is potentially mediated at least partly by factors other than viral load as estimated using Ct values. We found evidence that a model adjusting for a range of covariates could explain some of the population-level variation over time but systematically underestimated Ct values when incidence was increasing, and overestimated them when incidence was decreasing. This indicates the utility of Ct values from this data source as a tool for surveillance, potentially avoiding some of the biases of aggregated positive counts.

## Introduction

Since the start of the SARS-CoV-2 pandemic in late 2019, hundreds of millions of people have been confirmed as infected with the virus. Confirmation of infection has most commonly been done by reverse transcriptase polymerase chain reaction (RT-PCR), with test results indicated as positive if the cycle threshold (Ct) is below a certain level, negative if it is not, and sometimes as ambiguous in an intermediate range. However, the level of the cycle threshold (the Ct value) is a proxy of viral load of test-positive cases (Walker et al., 2021), which contains additional information on the infection. Viral loads can be an indicator of severity and transmissibility (Fajnzylber et al., 2020; Knudtzen et al., 2021; Lee et al., 2021; Lyngse et al., 2021). They vary widely between individuals (Challenger et al., 2022). Part of this variation can be explained by changes in viral load over the course of infection in individuals (Kissler, Fauver, Mack, Olesen, et al., 2021). The shape of this so-called viral load curve has been found to vary by age, infecting variant and vaccination status (Hay et al., 2022; Jones et al., 2021; Kissler, Fauver, Mack, Tai, et al., 2021). At the population level this can translate to changes in Ct values over time (Kissler, Fauver, Mack, Olesen, et al., 2021; Walker et al., 2021). These population-level changes have been shown potetnial as a surveillance tool as they allow better recovery of the underlying transmission process than aggregate counts of positive cases alone (Hay et al., 2021).

Because of these properties, there is value in continuing to characterise viral loads in those infected with SARS-CoV-2 and monitor changes therein with changing immunity status due to vaccination or prior infection, as well as with new emerging variants. Currently studies that seek to do this have largely been limited to non-surveillance datasets where multiple PCR results per person are available, cross-sectional studies, or simple summary statitics of Ct values generated via surveillance of those seeking tests. Here, we analyse Ct values as a proxy viral loads in more than 3 million symptomatic COVID-19 cases identified through routine testing in England since relatively early in the pandemic, as well as the degree to which individual Ct values can be predicted as a function of known individual characteristics such as age, vaccine status, and variant status. The main aim of this work is to further investigate the utility of Ct values for understanding transmission dynamics in real time in conjunction with other metrics such as case numbers.

## Methods

### Data

We used individual N gene Ct values from symptomatic pillar 2 cases (i.e., mostly from self-initiated testing from individuals presenting with symptoms) in England with symptom onset during or after the week beginning 2020-08-23 alongside data on the week of symptom onset, sex, source laboratory, age, reinfection status, and number of doses of vaccination received. We only included those with a positive test up to 6 days following symptom onset to avoid issues with recall bias. We limited the data set to tests performed using the Thermofisher TaqPath platform, in order to be able to assign variant status. We assigned variant status based on either genetic confirmation or, where no genotyping or sequencing result was available, timing and S-gene target failure (SGTF) status (Table 1). Observations with missing data for any of the extracted variables were dropped from further analysis. Combinations of number of vaccine doses and variants with fewer than 1000 observations were then dropped from the extracted data, leaving 3,706,330 unique cases with a Ct value (Table 2). We combined variant status, number of vaccines recived, and reinfection status (known reinfection or no record of prior infection) into a single composite variable to streamline the modelling process. We defined epidemic phase for each variant as months where they were clearly increasing or decreasing, such that the Alpha variant was defined to be increasing in December 2020 and decreasing in January 2021, the Delta variant was defined to be increasing in June 2021 and decreasing in December 2021, and the Omicron variant was defined to be increasing in December 2021 and decreasing in January 2022.

**Table 1:** Variant assignment for cases with S-gene result but without genetic confirmation.

| Variant | Condition |
| --- | --- |
| Wildtype | Specimen date < 2021-01-01 AND S-gene positive |
| Alpha | Specimen date < 2021-04-01 AND S-gene negative |
| Delta | 2021-05-01 <= Specimen date < 2022-01-01 AND S-gene positive |
| Omicron | Specimen date >= 2021-12-01 AND S-gene negative |
| Omicron BA.2 | Specimen date >= 2022-02-02 AND S-gene positive |

**Table 2:** Number of individual Ct values for each combination of variant and vaccine doses

| Variant | Vaccine doses | n |
| --- | --- | --- |
| Wildtype | 0 | 504,080 |
| Alpha | 0 | 514,831 |
| Alpha | 1 | 22,096 |
| Delta | 0 | 681,592 |
| Delta | 1 | 192,124 |
| Delta | 2 | 561,545 |
| Delta | 3 | 17,828 |
| Omicron BA.1 | 0 | 266,090 |
| Omicron BA.1 | 1 | 68,746 |
| Omicron BA.1 | 2 | 392,339 |
| Omicron BA.1 | 3 | 356,801 |
| Omicron BA.2 | 0 | 33,537 |
| Omicron BA.2 | 1 | 5,439 |
| Omicron BA.2 | 2 | 20,644 |
| Omicron BA.2 | 3 | 68,638 |
| Total |  | 3,706,330 |

### Model

We first fitted a simple generalised linear model with fixed effects for each day since onset, reinfection status, number of vaccine doses, and variant status. We assumed gamma distributed observations based on visual inspection of the underling data and the identification of a right skew with a moderately heavy tail. From this model we then estimated empirical mean Ct values for each interaction level along with 95% credible intervals.

We then modelled Ct values *y* for nucleocapsid protein (N) as target gene and assumed an additive relationship between covariates with gamma distributed errors using a generalised addititive modelling framework (Wood, 2011), again with gamma observation model and with the same error model across all covariates. Additive effects were used based on the assumption that individual covariates had independent effects on Ct value and hence viral load. We adjusted mean Ct values (*µ*) for sex, source laboratory, number of vaccines, variant status, and reinfection status (with number of vaccines, variant status, and reinfection status being treated as a composite variable as previously outlined) with fixed effects. We modelled time since symptom onset and age and the interaction of these effects smoothly using a 2-dimensional tensor product smooth (using a basis dimension of 7 and 10, respectively), estimated separately for each combination of variant, number of vaccine doses, and reinfection status (Wood, 2011). Mathematically this can be represented as follows:

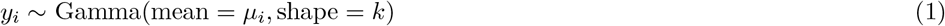

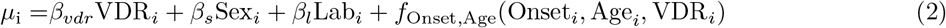

where VDR_*i*_ indicates the variant, number of vaccine doses received and reinfection status (known reinfection or no record of prior infection) of a given individual, Sex their sex, Lab laboratory where the Ct values where measured, and *f*_Onset,Age_ a 2-dimensional tensor product smooth varying as a function of days since symptom onset and age, estimated separately for each combination of variant, number of doses and reinfection status.

### Statistical inference

We report upper and lower bounds (based on a 95% credible interval) of the mean Ct values (based on plus/minus two standard deviations from the mean) as estimated using our simple generalised linear model, for each day since onset, number of vaccine doses, variant status, and reinfection status unadjusted for age or laboratory status. We then report predicted mean Ct values from our generalised additive model for each week along with their interquartile range compared to estimated reported mean Ct values as estimated using a model that treated each data stratification independently (along with the 95% credible interval) stratified by variant status. Alongside this we present case numbers per variant (limited to those included in this study) and estimated mean Ct values by days since onset, variant status, and epidemic phase (increasing or declining).

### Implementation

The model was implemented using the mgcv (Wood, 2011) package version 1.8-36 in R version 4.1.2 (R Core Team, 2019). All code and data required to reproduce this analysis is available from https://github.com/epiforecasts/covid19_ct_pillar2.

## Results

Mean viral loads as estimated from Ct values in the unvaccinated varied as a function of the number of days since symptom onset, with a maximum at one (Delta) or two (all other variants) days after infection (Figure 1A). Estimated viral loads at peak were highest for Delta with, however, an earlier decline as a function of time since symptom onset compared to Alpha, the variant with the second highest viral load. Estimated viral loads from Omicron infections declined slower as a function of time since symptom onset than from any of the other variants.

**Figure 1:**
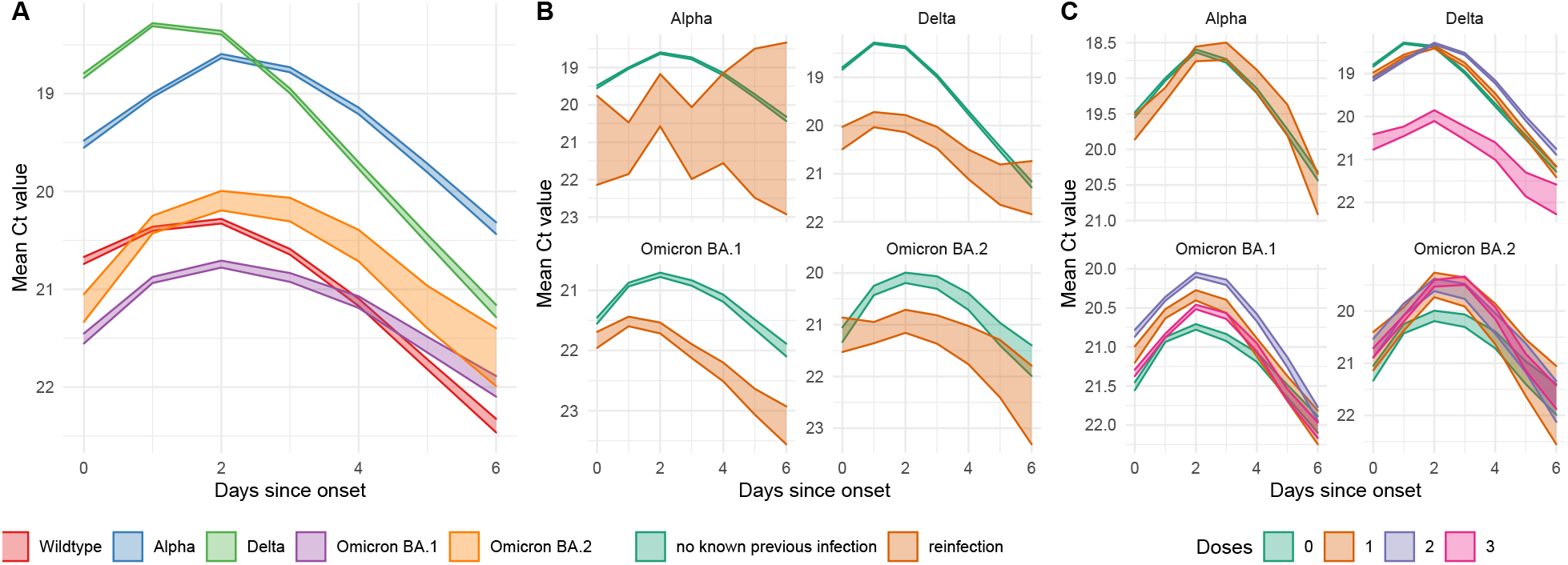
Estimated mean Ct values in (A) those unvaccinated without any known prior infection, in those unvaccinated as a function of whether there was knowledge of any prior infection (B), and in those without any known prior infection as a function of the number of vaccine doses (C). Bands indicate the 95% credible interval for the mean Ct value

With all variants, evidence of a prior infection reduced the estimated mean viral loads in those that with symptomatic infection (Figure 1B). The number of vaccine doses, on the other hand, had less of a discernible effect, with the most discernible effect a reduction in viral loads for those with three doses of vaccination and infected with Delta (Figure 1C).

Ct values changed over time, largely in response to a change in the dominant variant but also whilst one variant was dominating. Modelling Ct values as a function of time since symptom onset, age, evidence of prior infection and number of vaccine doses yields poor predictions at the individual level (deviance explained: 9.6%) yet broadly recovers the time-trends in mean Ct values (Figure 2A-B). That said, it appears to do so mostly by capturing the changes with different variants, whilst being less able to recover trends in Ct values whilst a variant is dominating. Part of this may be explained by epidemic phase bias, as symptom onset follows infection and therefore, during periods of exponentially increasing infections, is biased towards more recent infections Hay et al. (2021). This would imply that when the model underestimates Ct values for a given variant then infections are likely to be increasing and when it overestimates Ct values for a given variant they are likely to be decreasing. Indeed we observe higher estimated viral loads during periods of increase than during periods of decrease for all variants (Figure 2C). The observed trajectory of Ct values compared to the case trajectories also matches this interpretation of the mechanism between predicted and observed Ct values.

**Figure 2:**
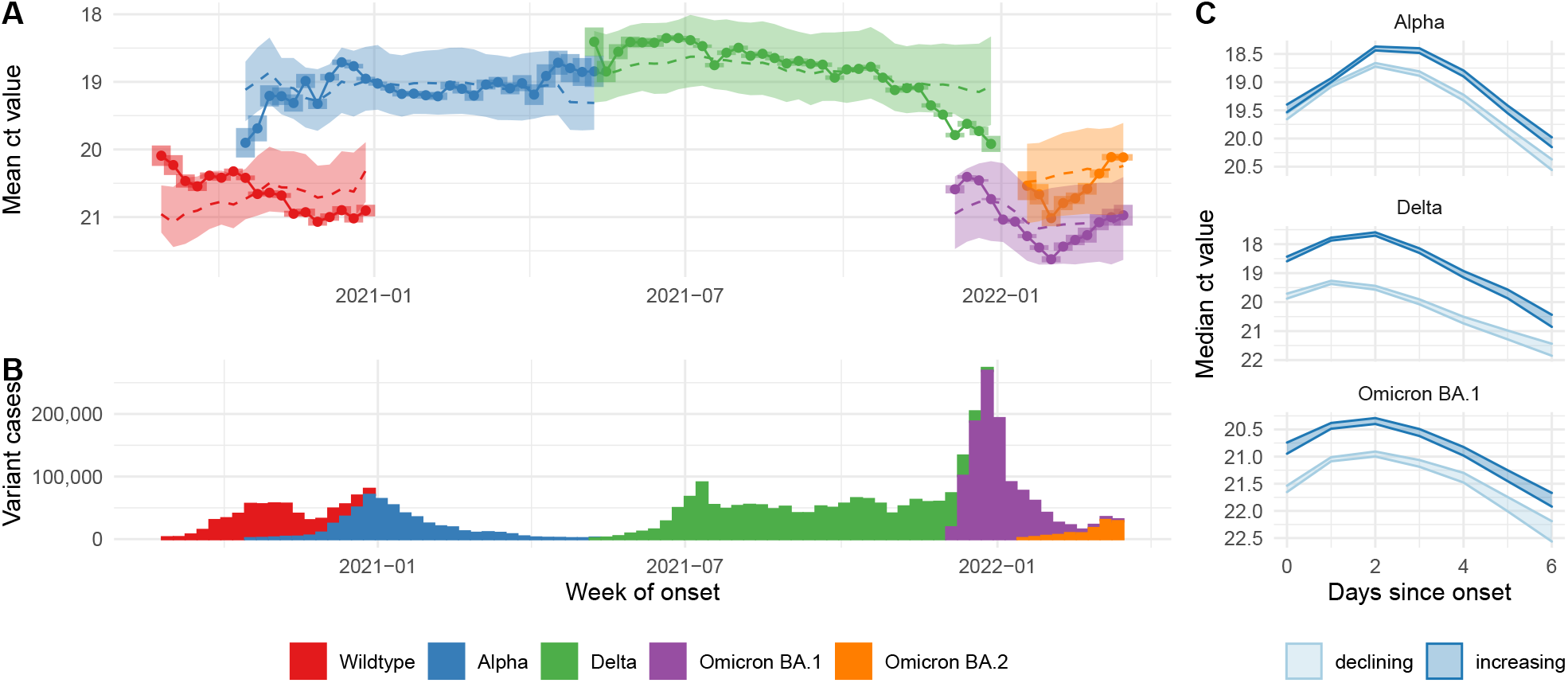
(A) Mean ct values (points and solid lines) estimated independently for each week and variant, with associated 90% confidence intervals as bars; means of modelled ct values (dashed lines) and their interquartile ranges (ribbons). (B) Case numbers identified as being due to a give variant. (C) Ct values by epidemic phase for three variants in unvaccinated people without known previous infection.

## Discussion

Analysing a large data set of Ct values in symptomatic COVID-19 cases in England, we found variation in Ct values (and hence viral loads by proxy) by time since symptom onset, vaccine and reinfection status, age and variant. We found highest viral loads in those tested 1-3 days after symptom onset for each of the variants, including when considering variation by vaccination or reinfection status. We further found that the Alpha and Delta variants produced higher viral loads than wildtype and Omicron variants, and that both vaccination and prior infection appeared to reduce viral loads.

In comparing these results to observations made with other data sets, it is important to bear in mind the specific biases that exist in the data we analysed. We only considered positive tests in symptomatic individuals as part of the so-called Pillar 2 testing in England, the vast majority of which would have been based on self-initiated testing at a community. Because of this, our data may preferentially reflect cases with symptoms that are both noticable enough to prompt individuals to get tested whilst not severe enough that the first test would happen in hospital, with age and other variables probably affecting the probability of getting tested as a function of viral load. Moreover, we did not have any information on why individuals would seek out a test only on day 5 or 6 of symptoms and whether this reflects a particular symptom trajectory such as from very mild to more severe, potentially with corresponding behaviour in the viral load trajectory. All that said, whilst these biases and the selection bias of Ct values less than 30 means our observed Ct values cannot be interpreted as samples of individual-level viral load trajectories as in previous work, their smooth behaviour which is broadly in line with that observed previously (Jones et al., 2021; Kissler, Fauver, Mack, Olesen, et al., 2021), indicates that conclusions on broad trends may be valid and should be further investigated.

It has previously been suggested that viral loads may be related to both severity and transmissibility, this does not appear to translate into a relationship that would make it possible to infer these directly and independently from observed viral loads. Of these two aspects viral loads in the data presented here appears more closely related to severity, previously observed to have been higher in Alpha and Delta than the previous or subsequent variants to date (Davies et al., 2021; Twohig et al., 2022), and lower than with Omicron (Nyberg et al., 2022), although the greater severity of Delta. The Omicron variant, on the other hand, which has shown a transmission advantage but decreased severity compared to other lineages (Nyberg et al., 2022; Pearson et al., 2021), shows significantly reduced viral loads. Combined with recent evidence that Ct values do not always correspond to infectious viral loads (Puhach et al., 2022), this suggests caution is warranted when trying to infer viral characteristics from Ct values.

It has further been suggested that Ct values may be a useful quantity to monitor as part of real-time surveillance of infectious disease dynamics (Hay et al., 2021; Lin et al., 2022; Tso et al., 2021). Our results, too, show evidence of “epidemic phase bias”, whereby a model taking into account relevant determinants of Ct values such as variant, age and variant and vaccination status overestimates Ct values at times of increasing case numbers and underestimates them at times of decreasing case numbers. That said, whilst this study adds to the evidence that Ct values can correspond to epidemic growth rate, we are not aware of any work that has demonstrated the use of a practical tool that can be used in real-time to improve predictions of infectious disease dynamics. Here, too, the real-time benefit is only suggested as we have used the full retrospective data set in all analyses rather than assessing the value of Ct values available in real time. There is clearly scope for further research into whether integrating Ct values from routine surveillance sources into analysis of real-time dynamics, in particular at a time when community may have been scaled back and overall case numbers are sampled from a smaller proportion of the population, could improve the accuracy and timeliness of estimates.

## Data Availability

The Pillar 2 surveillance data used in this repository is currently not publicly available. Access has been provided by the UK Health Security Agency (UKHSA) through the Scientific Pandemic Influenza Group on Modelling, Operational sub-group (SPI-M-O) for the Scientific Advisory Group for Emergencies (SAGE), where this work was first presented.

https://github.com/epiforecasts/covid19_ct_pillar2/

## Acknowledgements

We are grateful to the Scientific Pandemic Influenza Group on Modelling (SPI-M) for discussions of this work, and to the UK Health Security Agency (UKHSA) for sharing the data. We thank Sang Woo Park for helpful comments. SF and SA were funded by a Wellcome Trust Senior Research Fellowship (to SF, 210758/Z/18/Z).

## References

Challenger, J. D., Foo, C. Y., Wu, Y., Yan, A. W. C., Marjaneh, M. M., Liew, F., Thwaites, R. S., Okell, L. C., & Cunnington, A. J. (2022). Modelling upper respiratory viral load dynamics of SARS-CoV-2. BMC Medicine, 20 (1). https://doi.org/10.1186/s12916-021-02220-0

Davies, N. G., Jarvis, C. I., CMMID COVID-19 Working Group, Edmunds, W. J., Jewell, N. P., Diaz-Ordaz, K., & Keogh, R. H. (2021). Increased mortality in community-tested cases of SARS-CoV-2 lineage b.1.1.7. Nature, 593 (7858), 270–274. https://doi.org/10.1038/s41586-021-03426-1

Fajnzylber, J., Regan, J., Coxen, K., Corry, H., Wong, C., Rosenthal, A., Worrall, D., Giguel, F., Piechocka-Trocha, A., Atyeo, C., Fischinger, S., Chan, A., Flaherty, K. T., Hall, K., Dougan, M., Ryan, E. T., Gillespie, E., Chishti, R., Li, Y., … Li, J. Z. (2020). SARS-CoV-2 viral load is associated with increased disease severity and mortality. Nature Communications, 11 (1). https://doi.org/10.1038/s41467-020-19057-5

Hay, J. A., Kennedy-Shaffer, L., Kanjilal, S., Lennon, N. J., Gabriel, S. B., Lipsitch, M., & Mina, M. J. (2021). Estimating epidemiologic dynamics from cross-sectional viral load distributions. Science, 373 (6552). https://doi.org/10.1126/science.abh0635

Hay, J. A., Kissler, S. M., Fauver, J. R., Mack, C., Tai, C. G., Samant, R. M., Connelly, S., Anderson, D. J., Khullar, G., MacKay, M., Patel, M., Kelly, S., Manhertz, A., Eiter, I., Salgado, D., Baker, T., Howard, B., Dudley, J. T., Mason, C. E., … Grad, Y. H. (2022). Viral dynamics and duration of PCR positivity of the SARS-CoV-2 omicron variant. https://doi.org/10.1101/2022.01.13.22269257

Jones, T. C., Biele, G., Mühlemann, B., Veith, T., Schneider, J., Beheim-Schwarzbach, J., Bleicker, T., Tesch, J., Schmidt, M. L., Sander, L. E., Kurth, F., Menzel, P., Schwarzer, R., Zuchowski, M., Hofmann, J., Krumbholz, A., Stein, A., Edelmann, A., Corman, V. M., & Drosten, C. (2021). Estimating infectiousness throughout SARS-CoV-2 infection course. Science, 373 (6551). https://doi.org/10.1126/science.abi5273

Kissler, S. M., Fauver, J. R., Mack, C., Olesen, S. W., Tai, C., Shiue, K. Y., Kalinich, C. C., Jednak, S., Ott, I. M., Vogels, C. B. F., Wohlgemuth, J., Weisberger, J., DiFiori, J., Anderson, D. J., Mancell, J., Ho, D. D., Grubaugh, N. D., & Grad, Y. H. (2021). Viral dynamics of acute SARS-CoV-2 infection and applications to diagnostic and public health strategies. PLOS Biology, 19 (7), e3001333. https://doi.org/10.1371/journal.pbio.3001333

Kissler, S. M., Fauver, J. R., Mack, C., Tai, C. G., Breban, M. I., Watkins, A. E., Samant, R. M., Anderson, D. J., Metti, J., Khullar, G., Baits, R., MacKay, M., Salgado, D., Baker, T., Dudley, J. T., Mason, C. E., Ho, D. D., Grubaugh, N. D., & Grad, Y. H. (2021). Viral dynamics of SARS-CoV-2 variants in vaccinated and unvaccinated persons. New England Journal of Medicine, 385 (26), 2489–2491. https://doi.org/10.1056/nejmc2102507

Knudtzen, F. C., Jensen, T. G., Lindvig, S. O., Rasmussen, L. D., Madsen, L. W., Hoegh, S. V., Bek-Thomsen, M., Laursen, C. B., Nielsen, S. L., & Johansen, I. S. (2021). SARS-CoV-2 viral load as a predictor for disease severity in outpatients and hospitalised patients with COVID-19: A prospective cohort study. PLOS ONE, 16 (10), e0258421. https://doi.org/10.1371/journal.pone.0258421

Lee, L. Y., Rozmanowski, S., Pang, M., Charlett, A., Anderson, C., Hughes, G. J., Barnard, M., Peto, L., Vipond, R., Sienkiewicz, A., Hopkins, S., Bell, J., Crook, D. W., Gent, N., Walker, A. S., Peto, T. E., & Eyre, D. W. (2021). SARS-CoV-2 infectivity by viral load, s gene variants and demographic factors and the utility of lateral flow devices to prevent transmission. https://doi.org/10.1101/2021.03.31.21254687

Lin, Y., Yang, B., Cobey, S., Lau, E. H. Y., Adam, D. C., Wong, J. Y., Bond, H. S., Cheung, J. K., Ho, F., Gao, H., Ali, S. T., Leung, N. H. L., Tsang, T. K., Wu, P., Leung, G. M., & Cowling, B. J. (2022). Incorporating temporal distribution of population-level viral load enables real-time estimation of COVID-19 transmission. Nature Communications, 13 (1). https://doi.org/10.1038/s41467-022-28812-9

Lyngse, F. P., Mølbak, K., Træholt Franck, K., Nielsen, C., Skov, R. L., Voldstedlund, M., Cohen, A. S., & Kirkeby, C. T. (2021). Association between SARS-CoV-2 transmissibility, viral load, and age in households. https://doi.org/10.1101/2021.02.28.21252608

Nyberg, T., Ferguson, N. M., Nash, S. G., Webster, H. H., Flaxman, S., Andrews, N., Hinsley, W., Bernal, J. L., Kall, M., Bhatt, S., Blomquist, P., Zaidi, A., Volz, E., Aziz, N. A., Harman, K., Funk, S., Abbott, S., Hope, R., Charlett, A., … Thelwall, S. (2022). Comparative analysis of the risks of hospitalisation and death associated with SARS-CoV-2 omicron (b.1.1.529) and delta (b.1.617.2) variants in england: A cohort study. The Lancet. https://doi.org/10.1016/s0140-6736(22)00462-7

Pearson, C. A. B., Silal, S. P., Li, M. W. Z., Dushoff, J., Bolker, B. M., Abbott, S., Schalkwyk, C. van, Davies, N. G., Barnard, R. C., Edmunds, W. J., Bingham, J., Meyer-Rath, G., Jamieson, L., Glass, A., Wolter, N., Govender, N., Stevens, W. S., Scott, L., Mlisana, K., … Pulliam, J. R. C. (2021). Bounding the levels of transmissibility & immune evasion of the omicron variant in south africa. https://doi.org/10.1101/2021.12.19.21268038

Puhach, O., Adea, K., Hulo, N., Sattonnet, P., Genecand, C., Iten, A., Bausch, F. J., Kaiser, L., Vetter, P., Eckerle, I., & Meyer, B. (2022). Infectious viral load in unvaccinated and vaccinated individuals infected with ancestral, delta or omicron SARS-CoV-2. Nature Medicine. https://doi.org/10.1038/s41591-022-01816-0

R Core Team. (2019). R: A language and environment for statistical computing. R Foundation for Statistical Computing. https://www.R-project.org/

Seaman, S. R., Nyberg, T., Overton, C. E., Pascall, D., Presanis, A. M., & De Angelis, D. (2021). Adjusting for time of infection or positive test when estimating the risk of a post-infection outcome in an epidemic. https://doi.org/10.1101/2021.08.13.21262014

Tso, C. F., Garikipati, A., Green-Saxena, A., Mao, Q., & Das, R. (2021). Correlation of population SARS-CoV-2 cycle threshold values to local disease dynamics: An exploratory observational study (preprint). JMIR Public Health and Surveillance. https://doi.org/10.2196/28265

Twohig, K. A., Nyberg, T., Zaidi, A., Thelwall, S., Sinnathamby, M. A., Aliabadi, S., Seaman, S. R., Harris, R. J., Hope, R., Lopez-Bernal, J., Gallagher, E., Charlett, A., De Angelis, D., Presanis, A. M., Dabrera, G., & the COVID-19 Genomics UK (COG-UK) consortium. (2022). Hospital admission and emergency care attendance risk for SARS-CoV-2 delta (b.1.617.2) compared with alpha (b.1.1.7) variants of concern: A cohort study. The Lancet Infectious Diseases, 22 (1), 35–42. https://doi.org/10.1016/s1473-3099(21)00475-8

Walker, A. S., Pritchard, E., House, T., Robotham, J. V., Birrell, P. J., Bell, I., Bell, J. I., Newton, J.N., Farrar, J., Diamond, I., Studley, R., Hay, J., Vihta, K.-D., Peto, T. E., Stoesser, N., Matthews, P. C., Eyre, D. W., & Pouwels, K. B. (2021). Ct threshold values, a proxy for viral load in community SARS-CoV-2 cases, demonstrate wide variation across populations and over time. eLife, 10. https://doi.org/10.7554/elife.64683

Wood, S. N. (2011). Fast stable restricted maximum likelihood and marginal likelihood estimation of semiparametric generalized linear models. Journal of the Royal Statistical Society (B), 73 (1), 3–36.

